# Are CNV Risk Scores Linked to Neurodevelopmental and Mental Health Characteristics Within CNV-Associated Intellectual Disability?

**DOI:** 10.64898/2026.07.14.26358034

**Authors:** Zhaotian Chi, Aaron Alexander-Bloch, Sharon AS Neufeld, Jeanne Wolstencroft, David Skuse, IMAGINE-ID consortium, Kate Baker

**Affiliations:** MRC Cognition and Brain Sciences Unit, University of Cambridge, U.K; Department of Psychiatry, University of Cambridge, U.K; Department of Psychiatry, University of Pennsylvania, Philadelphia, U.S.A; Department of Child and Adolescent Psychiatry and Behavioral Science, Children’s Hospital of Philadelphia, Philadelphia, U.S.A; Great Ormond Street Institute of Child Health, University College London, U.K; Department of Pathology, University of Cambridge, U.K

**Keywords:** Intellectual disability, copy number variants, neurodevelopmental disorders, mental health, genomics

## Abstract

**Background:** Children and young people (CYP) with intellectual disability (ID) frequently have co-occurring neurodevelopmental (ND) and mental health (MH) difficulties. While copy number variants (CNVs) are identified as an important aetiology of ID, it is unclear whether and how CNV risk scores predict ND and MH characteristics within the CNV-associated ID population.

**Methods:** We analysed data from the UK-based IMAGINE-ID cohort of CYP (aged 4–19 years) with ID and clinically-reported CNVs (N = 1,640). CNVs were annotated with Gencode 19 in ENSEMBL to calculate CNV risk scores, including summed probability of loss-of-function intolerance (pLI) and dosage sensitivity. Multivariate regression models examined the prediction of CNV variables and inheritance on ND and MH characteristics, assessed via the Development and Well-Being Assessment (DAWBA). Post-hoc analyses explored CNV variable stratification (lower vs. higher range pLI).

**Results:** Higher summed pLI scores (indexing CNV genes’ intolerance to loss of function) unexpectedly predicted fewer MH difficulties and a lower likelihood of ND diagnoses, even after accounting for demographic factors and CNV inheritance. Post-hoc analyses identified a threshold effect. Within the lower pLI range, higher pLI scores were associated with greater MH difficulties, consistent with findings from population-based samples. In contrast, within the higher pLI range, higher pLI scores were associated with fewer MH difficulties (among individuals more likely to have severe ID).

**Conclusion:** These findings challenge the assumption that CNV genomic “risk scores” universally predict ND and MH difficulties. Instead, within CNV-associated ID, complex relationships exist between CNV risk scores, inheritance and phenotypes. These insights emphasise the necessity of integrating genomic results with familial and developmental context to understand individual vulnerabilities and support needs.

## Introduction

Intellectual disability (ID) is defined by significant impairments in cognitive abilities and adaptive functioning, reflecting early-onset differences in brain development, with lifelong impacts on independence and social integration.^1^ Children and young people (CYP) with ID can display other neurodevelopmental (ND) difficulties, such as attention deficit/hyperactivity disorder (ADHD) and autism spectrum disorder (ASD).^2,3^ Moreover, they also experience elevated rates of mental health (MH) symptoms such as conduct disorders, anxiety and depression. Meta-analytic findings suggest that 30–50% of CYP with ID meet diagnostic criteria for at least one psychiatric disorder, compared with 8–18% in non-ID groups.^4,5^ However, not all CYP with ID experience ND or MH difficulties, and understanding the diverse factors influencing co-occurrence may help to explain heterogeneity within this population and inform targeted support strategies.^6–8^

While demographic and medical factors such as age, gender, socioeconomic status (SES), and physical illness have been linked to MH variation within ID populations, the contribution of genetic factors remains under-explored.^9,10^ Advances in genomic sequencing technologies have enabled the identification of causal or contributory variants in an increasing proportion of people with ID,^11–13^ opening new opportunities to investigate genetic influences on co-occurring MH and ND risks.^1,14^ Copy number variants (CNVs), defined as abnormal DNA dosage (most often deletions or duplications), have emerged as an important class of genetic aetiology of ID. Although individually rare or ultra-rare, CNVs collectively account for a substantial proportion of ID and associated conditions, and have become a focus of research and clinical application.^15^

There is accumulating evidence that CNVs contribute significantly to ND and MH traits, both within clinical samples and the general population.^16,17^ Within ID of known genetic aetiology, CNVs have been associated with a higher risk of emotional and behavioural problems, compared to other variant types such as single nucleotide variants (SNVs).^18^ Moreover, Chawner et al^19^ examined phenotypic profiles of children with 13 recurrent NDD-associated CNVs and sibling controls, and found that carrying any of these CNVs is linked with broad impairment on cognitive and mental health traits, including IQ, ADHD, ASD, and emotional and behavioural problems. Quantitative and qualitative differences between the 13 CNVs were found, predicting a moderate level of variance in IQ (18.5%), ADHD (14.5%), and conduct problems (17.5%). Hence, while CNVs may exert quantitatively and qualitatively specific influence on ND and MH, there is also extensive variation within CNV groups. CNV inheritance and socioeconomic status also contribute to variance – familial CNVs have been associated with elevated risks of emotional and behavioural problems compared with de novo or unknown origin CNVs, but these MH problems are also more likely to be observed in deprived social contexts and thus in combination with additional risk factors for MH problems more generally.^14^

An important question remains whether and how CNVs’ genomic properties may predict variation in ND and MH traits (alone, or in combination with CNV inheritance). This is especially important for rare (rather than recurrent) CNVs – which can occur anywhere across the genome and vary substantially in size and gene content – as they cannot be studied individually. Over the past decade, approaches have been developed to quantify the genomic impacts of recurrent and non-recurrent CNVs.^20,21^ CNVs are annotated based on diverse genomic properties to derive CNV risk scores, which have shown significant utility in predicting ND and MH traits across different study populations (see Table 1).

**Table 1.** CNV risk scores and previously-observed phenotype associations.

| ***Risk score index*** | ***Definition*** | ***Associations in general population samples*** | ***Associations in clinical samples*** |
| --- | --- | --- | --- |
| *CNV burden* | Number of genes or total base pairs affected | Lower IQ, attention difficulties, ASD traits (Auwerx et al., 2022; Guyatt et al., 2018) | “Second hit” CNV links to cognitive impairment and ND/MH difficulties (Pizzo et al., 2019) |
| *Loss-of-function intolerance: pLI (probability of loss-of-function intolerance)* | A gene’s sensitivity to disruptive mutations (LoF constraint), derived from large-scale population sequencing data (Lek et al., 2016) | Reduced cognitive ability and increased ND/MH symptoms (Huguet et al., 2018) | Predicted reduced general intelligence and increased autism risk within ASD samples (Douard et al., 2021) |
| *Dosage sensitivity: pHI (probability of haploinsufficiency), pTS (probability of triplosensitivity)* | A gene’s dosage sensitivity based on its contribution to multiple disorders derived from meta-analyses of rare CNVs (Collins et al., 2022) | Both pTS and pHI predicted cognitive functions, externalising symptoms, and psychosis-spectrum symptoms (Alexander-Bloch et al., 2022) | No known studies. |

One recent study combined and compared multiple CNV risk scores to assess their impact on MH and cognitive functions within the Philadelphia Neurodevelopmental Cohort (PNC), a large community-based sample of youth recruited through a paediatric healthcare network.^22^ CNV risk scores were highly inter-correlated, and predicted variation within cognitive and psychopathological domains. Specifically, deletion pHI scores were associated with cognitive accuracy, externalising problems, and categorical psychiatric diagnoses (psychosis-spectrum disorder and ADHD, but not anxiety or depression).

In summary, while previous studies have applied CNV risk scores to predict developmental outcomes in paediatric populations, they have not been examined in individuals with CNV-associated ID. The primary aim of this study was to examine associations between CNV risk scores and ND and MH outcomes in CYP with CNV-associated ID. We focused on participants with a single deletion or duplication in the main analysis, while including those with multiple CNVs in supplementary analyses. A secondary aim was to investigate whether associations between CNV risk scores and ND/MH outcomes differed according to CNV inheritance. Finally, following the initial analyses, we explored a post-hoc question: whether genomic–ND/MH associations differed in direction or magnitude across subgroups defined by lower-range (comparable to population-based samples) versus higher-range CNV risk scores.

## Methods

### Sample

This study analysed data from the IMAGINE-ID study (Intellectual Disability and Mental Health: Assessing the Genomic Impact on Neurodevelopment), a UK-based cohort of 3,407 children with developmental delay or ID, each with a molecular genetic diagnosis.^14^ Ethics approval for the IMAGINE-ID study was obtained from the London–Queen Square Research Ethics Committee.

Within the IMAGINE-ID cohort, 1,646 children aged 3 to 19 years were diagnosed with CNVs. For the main analyses, we selected participants with a single CNV (n = 1,384; 849 deletions, 535 duplications), and conducted analyses separately for deletions and duplications. In the extended analysis, we included all participants with either a single or multiple CNVs (total n = 1,640; including an additional n = 256 with multiple CNVs; CNV combination types are listed in Table S1).

### CNV risk score calculations

CNV data (size, chromosomal location, type, inheritance) were obtained from diagnostic genomic reports.^14^ Annotations were performed using Gencode V19 via the ENSEMBL genome browser, following the methods outlined in Alexander-Bloch et al.^22^ Additional details on the calculation procedures are provided in Supplementary Note 1. CNV risk scores included in the analysis were CNV burden (size in base pairs), summed loss-of-function intolerance for all encountered genes (pLI, LOEUF), and summed dosage sensitivity (pHaplo for deletions, pTriplo for duplications). The intolerance and dosage sensitivity indices of all encountered genes were derived from ENSEMBL genome browser and the supplementary materials of Collins et al,^20^ respectively. Inheritance type was categorised as de novo, familial or unknown, and dummy coded with the de novo group defined as the reference (coded 0).

For the extended analysis, we calculated participant-level risk scores (total numbers of CNVs, and a sum pLI score for all CNVs), and the sample was analysed collectively. Because of the complexity of multiple inheritance possibilities for individuals with multiple CNVs, the inheritance type was recoded as a binary variable, indicating whether any CNV was de novo (coded 1).

### Outcome measures

Primary caregivers completed online questionnaires about their child’s MH and ND characteristics. MH characteristics were assessed via the Development and Well-Being Assessment (DAWBA),^23^ yielding 11 algorithm-derived likelihood bands for ND and MH conditions (see Table S2). Scores ranged from 0 to 4 or 5, corresponding to increasing likelihood of the condition(<0.1% to >70%.^24^ The DAWBA has been widely used in CYP with neurodevelopmental conditions and was suggested to be generally applicable in ID groups.^14,25^

We derived a composite measure of MH difficulties (MH_sum) by summing nine DAWBA likelihood scores: separation anxiety, specific phobia, social phobia, obsessive–compulsive disorder (OCD), general anxiety, depression, oppositional defiant disorder (ODD), conduct disorder, and tic disorders (range: 0–41), to provide an index of overall psychiatric burden.

Children’s ND characteristics were reported using three indicators. ID severity was approximated by the developmental quotient (DQ), calculated as parent-reported “mental age” divided by chronological age. Physical disabilities were summarised by totalling eight binary DAWBA items (range: 0–8; see Table S3). ASD and ADHD likelihood scores from the DAWBA (both on 0–5 scales) were retained as separate outcomes because they represent distinct neurodevelopmental traits and are key phenotypes associated with ID.

### Data analyses

Data were managed and analysed in R using packages for genomic data analysis (BioManager)^27^, linear regression (lme4)^28^, and data visualisation (ggplot2)^29^.

We applied multivariate linear regression (MLR) to examine the predictive effects of CNV risk scores and inheritance type. Model comparisons were based on adjusted R² and Akaike Information Criterion (AIC) values, with higher R² and lower AIC indicating better model fit. Missing data were handled using pairwise deletion.^30^ All models adjusted for child age, gender, and socioeconomic status (SES), indexed by the Index of Multiple Deprivation (IMD) decile score (1 = most deprived).^26^

Given the high inter-correlation among CNV risk metrics, each risk score was first evaluated separately in covariate-adjusted univariate models. The CNV metric demonstrating the best overall model fit was selected for inclusion in subsequent multivariate models to minimise multicollinearity. MLR models were then conducted separately for deletions and duplications, regressing each outcome on demographic covariates, pLI, and CNV inheritance using a forward stepwise approach.

Following the primary analyses, we conducted an exploratory post hoc analysis to examine whether associations differed across the distribution of pLI scores. As the mean pLI in the IMAGINE-ID sample (M = 4.05, SD = 5.07) was substantially higher than that reported in a population-based cohort (M = 0.1, SD = 0.54)^22^, we stratified the sample at pLI = 1.18 (two standard deviations above the population mean). Regression models were then estimated separately in lower- (n=387) and higher-range (n=997) pLI subgroups.

## Results

### Sample Demographics and CNV characteristics

Descriptive statistics for the sample demographics and CNV characteristics are presented in Table 2. The mean age was approximately 8.5 years, with boys comprising around 59% of participants. The IMD decile score (range 1–10), was broadly distributed across the sample (M = 5.15, SD = 2.88), indicating representation across different socioeconomic ranges. Approximately 30% of the cohort (n = 415) carried one of the recurrent CNVs listed in Supplementary Table 4, with the remainder comprising unique or ultra-rare non-recurrent CNVs.

**Table 2.**
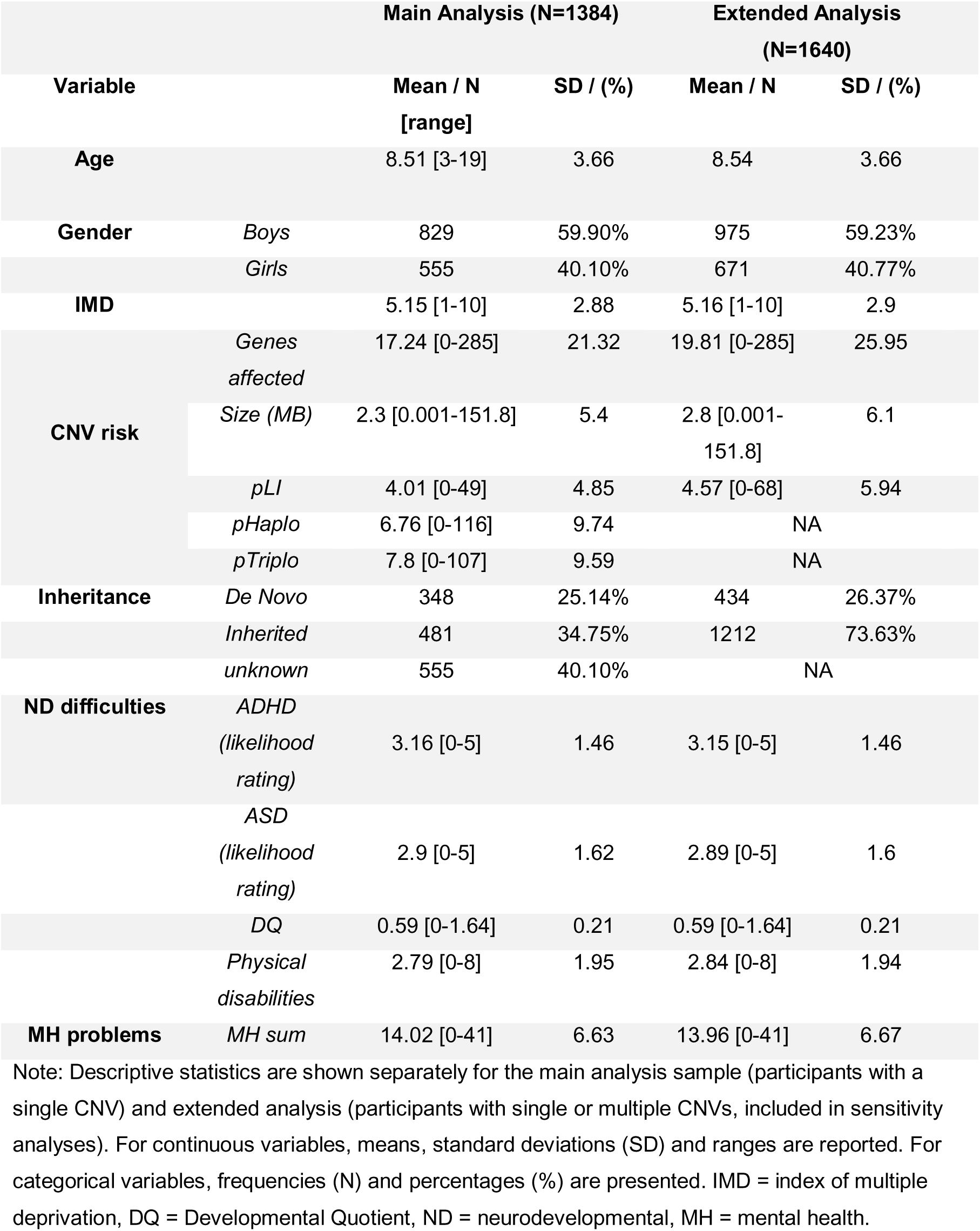
Summary of Descriptive Information.

### Correlation matrix

The correlation matrices of the main study variables are shown in Figure S3. CNV risk scores were highly inter-correlated. Surprisingly, summed pLI showed a negative correlation with MH, ADHD and ASD outcomes (r = –.09 to –.16). IMD was positively correlated with CNV risk scores, indicating greater socioeconomic affluence was linked to higher CNV risk score. Conversely, IMD was negatively correlated with MH scores, suggesting that higher affluence corresponded to lower MH risk. IMD was also associated with CNV inheritance, with inherited CNVs being more common among families from deprived backgrounds. CNV risk scores were moderately correlated with inheritance (ie, inherited CNVs tended to have lower pLI scores). In summary, correlation matrices highlight potential associations between CNV risk scores and ND/MH variables, though these associations are complex and may be confounded by SES and CNV inheritance effects.

### Comparison between CNV risk indices

To assess the predictive effect of different CNV variables, we first conducted univariate regression models with pLI, pHaplo/pTriplo, and CNV size as predictors, while controlling for the covariates (Table S5). Significant associations were observed between pLI and MH Sum, ASD, and ADHD scores for deletions, and between pLI and MH Sum for duplications. Dosage sensitivity scores (pHaplo/pTriplo) also showed associations with MH Sum. CNV size predicted physical disabilities and DQ. pLI outperformed other risk scores, as measured by the AIC, for predicting MH, ASD and ADHD scores, while CNV size best predicted DQ and physical disabilities. In view of inter-correlation between CNV risk indices, we selected pLI as a consistent CNV risk score for the subsequent analyses.

### Do CNV risk scores predict ND and MH outcomes?

We examined whether genomic features (pLI and inheritance) predict variation in MH and ND outcomes. Model comparisons (Table S6) in a stepwise inclusion method identified the best-fit models for each outcome. For deletions, models including covariates, pLI and inheritance best predicted all outcomes. For duplications, MH sum and ADHD were predicted by covariates, pLI and inheritance, whereas ASD, DQ and physical disabilities were best explained by covariates alone.

Model estimates are presented within Table 3. Within deletions, higher pLI was associated with lower risk of MH sum, ASD, and ADHD (β = −.11 to −.13, *p* < .004). Within duplications, higher pLI predicted lower MH sum (β = –.11, *p* = .03). Within deletions, de novo inheritance was linked to lower risk for MH and ADHD symptoms (β = 0.10 to 0.22, *p* < .03). However, de novo CNVs were also associated with more severe developmental delay i.e. lower DQ (familial vs. de novo: β = .15, *p* = .002; unknown vs. de novo: β = .16, *p* < .001) and greater physical disability (unknown vs. de novo: β = −.10, *p* = .03). Within duplications, de novo inheritance predicted lower risk of ADHD (β = .15, *p* = .03). Results for other outcomes were not interpreted due to poor model fit.

**Table 3.** Model estimates of the best-fit models for all outcomes.

| DV | IV | Deletions |  |  |  | Duplications |  |  |  |
| --- | --- | --- | --- | --- | --- | --- | --- | --- | --- |
| | | std $\beta$ | 95% CI lower | upper | p | std $\beta$ | 95%CI lower | upper | p |
| **MH_Sum** | pLI | -0.12 | -0.21 | -0.04 | **0.004** | -0.11 | -0.21 | -0.01 | **0.03** |
|  | Familial Inheritance | 0.22 | 0.12 | 0.31 | **<.001** | 0.14 | -0.01 | 0.3 | 0.07 |
|  | Unknown Inheritance | 0.18 | 0.09 | 0.27 | **<.001** | 0.09 | 0.06 | 0.24 | 0.23 |
| **ADHD** | pLI | -0.11 | -0.19 | -0.04 | **0.004** | -0.09 | -0.18 | 0.004 | 0.06 |
|  | Familial Inheritance | 0.1 | 0.01 | 0.19 | **0.03** | 0.03 | -0.12 | 0.17 | 0.7 |
|  | Unknown Inheritance | 0.13 | 0.04 | 0.21 | **0.003** | 0.15 | 0.02 | 0.29 | **0.03** |
| **ASD** | pLI | -0.13 | -0.2 | -0.05 | **<.001** |  |  |  |  |
|  | Familial Inheritance | -0.06 | 0.15 | 0.03 | 0.19 |  |  |  |  |
|  | Unknown Inheritance | -0.003 | -0.09 | 0.08 | 0.95 |  |  |  |  |
| **DQ** | pLI | 0.03 | -0.05 | 0.14 | 0.44 |  |  |  |  |
|  | Familial Inheritance | 0.15 | 0.06 | 0.28 | **0.002** |  |  |  |  |
|  | Unknown Inheritance | 0.16 | 0.07 | 0.27 | **<.001** |  |  |  |  |
| **Physical disabilities** | pLI | 0.04 | -0.04 | 0.12 | 0.38 |  |  |  |  |
|  | Familial Inheritance | -0.08 | -0.17 | 0.02 | 0.11 |  |  |  |  |
|  | Unknown Inheritance | -0.1 | -0.18 | 0.01 | **0.03** |  |  |  |  |

**Note:** significant effects are shown in bold. Inheritance was dummy coded with the de novo group as the reference category. MH\_Sum refers to the sum psychiatric likelihood score. Lower DQ and higher Physical Disability scores indicate higher levels of impairment. Results for ASD, DQ, and Physical Disability outcomes within duplications are not shown because covariate-only models provided the best fit. DV = dependent variable, IV = independent variable.

To summarise, higher pLI predicted fewer MH problems within both deletion and duplication subgroups, as well as a lower risk of ASD and ADHD for deletions. De novo CNVs were associated with a lower risk for MH problems for both deletions and duplications, but more severe developmental impairments for deletions only.

### Do effects of CNV risk scores on ND and MH outcomes differ according to CNV inheritance?

To test whether the effects of pLI differ within CNV inheritance groups, we conducted regression analyses separately within the de novo, inherited, and unknown inheritance groups. Full results are shown in Table S7. As no statistically significant associations were observed within duplication subgroups (*p* > .06), Figure 1 presents results for deletions only for clarity. Within the deletion subgroup, pLI negatively predicted MH, ASD, and ADHD scores for inherited and unknown CNVs (β = –.15 to –.29, *p* < .03), but not within the de novo group (*p* > .13). Within duplications, no significant associations between pLI and any outcome were detected for any inheritance group. These findings suggest that pLI effects may depend on CNV inheritance, particularly for deletions, although larger samples are required to confirm specificity.

**Figure 1:**
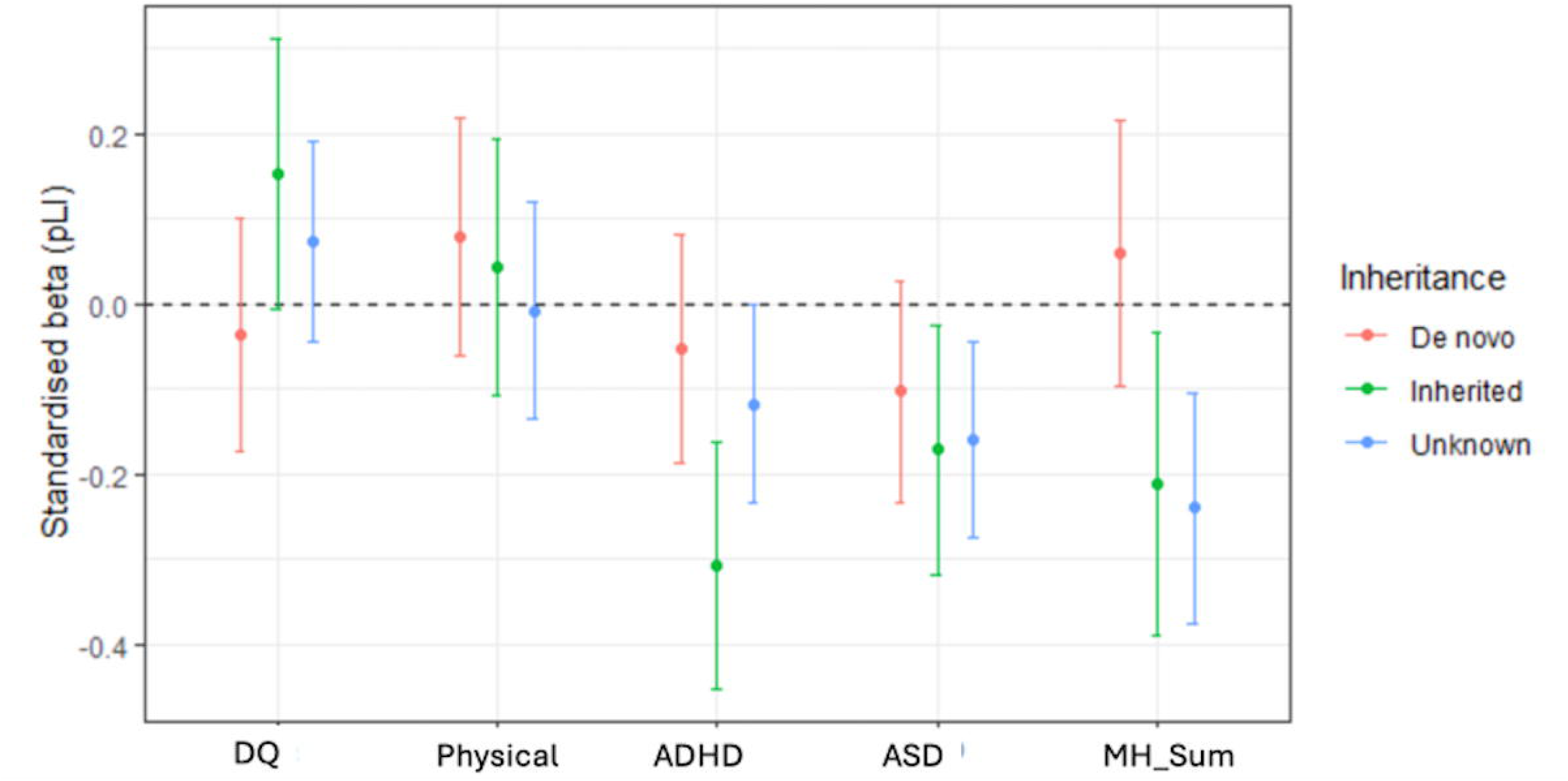
Association between pLI and outcomes (stratified by CNV inheritance) Effect sizes of pLI by inheritance group within deletion subgroup. Points represent standardised regression coefficients (β) for pLI, and error bars indicate 95% confidence intervals. Models were adjusted for age, sex, and socioeconomic status. Sample sizes were: de novo (n = 268), inherited (n = 232), and unknown (n = 349). Results for duplications are presented in Table S7 but are not visualised due to the absence of significant effects.

### Do effects of CNV risk scores on ND and MH outcomes differ according to pLI range?

The direction of effect of pLI observed in the above analysis seemed to contradict previous findings in a community-based sample ^22^. Notably, the distribution of summed pLI scores differed between the two samples, with pLI in the present study being higher, raising the possibility that associations may not be uniform across the full distribution of LoF-intolerance across the genome. To examine whether associations between CNV risk scores and ND and MH outcomes differed across levels of cumulative pLI burden, we explored whether associations between pLI and outcomes demonstrate a threshold effect. We stratified the sample by lower vs. higher pLI range (cut-off = 1.18). Subgroup analyses based on pLI ranges are shown in Figure 2 (more details are available in Supplementary Table S8 & S9).

**Figure 2:**
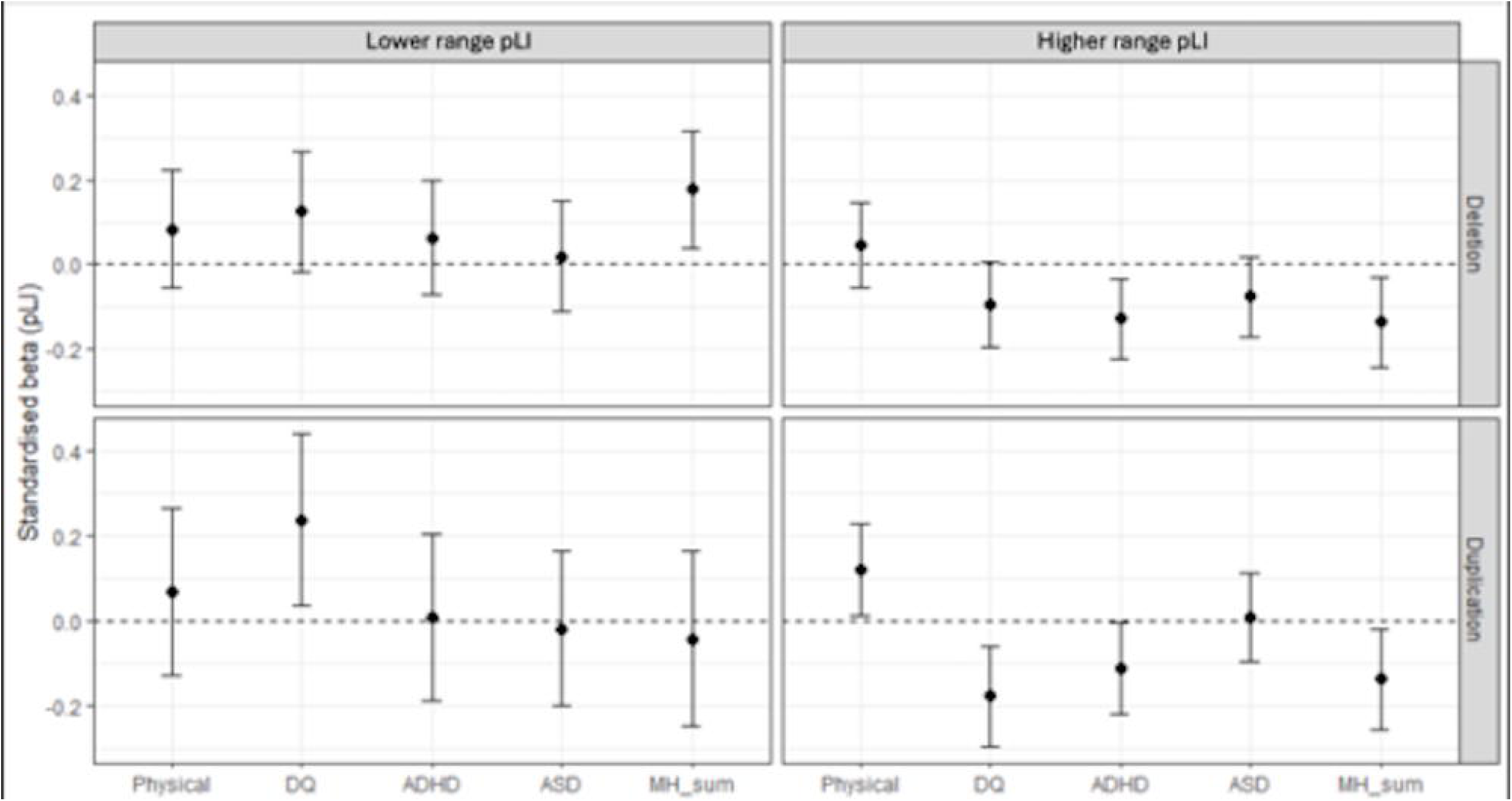
Association between pLI and outcomes (stratified by pLI range) Associations between pLI scores and neurodevelopmental and mental health outcomes stratified by pLI range and CNV type. Points represent standardised regression coefficients (β) for pLI from multivariable linear regression models. Error bars indicate 95% confidence intervals. The dashed horizontal line represents β = 0. Models were adjusted for age, sex, socioeconomic status (IMD decile), and CNV inheritance (familial and unknown vs. de novo reference). Panels are stratified by pLI range (Low ≤ 1.18; High > 1.18) and CNV type (deletions and duplications).

Within the deletion group with lower range pLI scores (≤ 1.18), higher pLI predicted higher MH likelihood score (β = .18, *p* = .01), consistent with previous findings in a paediatric community sample in which summed CNV loss-of-intolerance was associated with MH vulnerabilities.^22^ No significant effects of pLI were found for ASD, ADHD, DQ or physical disabilities within this subgroup. For deletions with higher pLI scores (pLI > 1.18), higher pLI predicted lower MH and ADHD scores (β = −.13, *p* < .01), as observed in the combined sample. No significant associations were found for ASD, DQ or physical disabilities.

Within the duplication subgroup, the effect of pLI also differed by pLI range. For lower-range pLI duplications, higher pLI was associated with lower impairment in DQ (β = .24, *p* = .01), but showed no significant associations with any other outcome. In contrast, in the higher range pLI duplication group, higher pLI predicted greater impairments in DQ (β = −.18, *p* = .003) and physical disability (β = .12, *p* = .03), but a lower risk for MH (β = −.14, *p* = .02) and ADHD problems (β = −.11, *p* = .04).

### Results of the extended analysis of multiple CNVs

Findings from the extended analyses, which included children with multiple CNVs, are presented in Table S1 and Tables S10-S13. Repeating sample-wide analysis of CNV risk scores as predictors of ND and MH variation, we again observed that higher pLI scores were generally associated with lower risks of MH and ADHD conditions. Analysing pLI-outcome relationships according to CNV inheritance status, the predictive effect of pLI on MH and ADHD was only evident in children without known de novo CNVs (i.e. where all CNVs were inherited or of unknown status). Finally, stratifying by summed pLI scores, the prediction of pLI is again different: in the lower range pLI group, higher pLI predicted higher DQ scores, whereas in the higher range pLI group, higher pLI predicted lower DQ scores. In contrast, inheritance status showed a more consistent pattern, with the presence of de novo CNV being associated with lower MH and ADHD likelihood.

## Discussion

Although genomic diagnosis is increasingly available within clinical assessment of individuals with ID, the relationships between specific genomic variants and each individual’s constellation of developmental difficulties are not fully understood. This study aimed to expand our understanding of genomic contributions to variation in ND and MH phenotypes, within the diverse population of children and young people with CNV-associated ID. We investigated the predictive value of CNV risk scores and CNV inheritance on ND and MH characteristics, within a large CNV-associated ID research cohort. We found that, in the context of ID and the presence of clinically-reported CNVs (presumed causal or contributory), relationships between risk scores and ND / MH phenotypes are present, but they are complex and can be counter-intuitive.

We first compared several CNV risk scores with respect to their predictive effects. We found that summed pLI (a measure of each CNV’s combined genetic content’s intolerance to disruption, derived from population-wide background variation) outperformed other risk score metrics in predicting MH and ND variables. At the whole cohort level, higher summed pLI predicted lower likelihoods of ND and MH conditions in children and young people with ID. Comparatively, Alexander-Bloch and colleagues^22^ found dosage sensitivity scores (pHaplo and pTriplo, derived from CNV analysis within disorder cohorts) were the best predictor of variation. This contrast may be explained by pLI distributions across cohorts – in the community-based sample, pLI was highly concentrated at a lower range (mean = 0.1, SD = 0.54), indicating lower risk, limited variance, and some outlier values. In contrast, pLI values in IMAGINE-ID were higher and showed greater variance, which may increase the power to detect associations with ND and MH outcomes in ID samples.

We then went on to model the contribution of CNV risk scores to within-cohort variation in MH and ND outcomes, taking demographic factors into account. We found that summed pLI for single deletions was predictive of lower MH, ASD and ADHD risks, and for single duplications pLI was predictive of lower MH risk. Model estimates in the larger expanded sample, combining pLI for multiple CNVs and analysing at participant-level irrespective of deletion or duplication, were broadly consistent with the primary analyses. PLI was associated with reduced risk of MH and ADHD diagnosis, but increased physical disabilities. Negative relationships between pLI and ND / MH risks are contrary to previous studies,^22,31^ and counter-intuitive, as higher pLI scores are expected to reflect greater biological vulnerability. One possibility is that CNV risks may influence cognitive development not adequately captured by the DQ and physical disability measures available in this dataset, which may counter-balance effects on mental health within an ID sample. Individuals with more severe cognitive impairments may show a relative reduction in co-occurring MH and ND difficulties, either because severe cognitive impairment constrains the manifestation of these difficulties, or because severe cognitive impairments overshadow the assessment and recognition of such problems.^32^ A linked explanation for this paradox lies in the neurodevelopmental gradient theory,^33^ which views CNV-associated risks as lying on a neurodevelopmental continuum, with a sequence of increasing impairment from bipolar disorders to ADHD, ASD, and ID. In this sense, more disruptive (higher pLI) CNVs might result in more severe ID and reduce the relative risk of developing ASD or ADHD, which are closer to typical development on this continuum i.e. lower pLI CNVs are more likely to lead to milder ID but greater risk of ND and MH difficulties.

On the other hand, this counter-intuitive result might also be a product of the complex inter-relationships between CNV risk, CNV inheritance and SES. Our correlation results indicate that higher SES is associated with higher pLI, as well as a higher likelihood for a de novo CNV, compared with inherited CNV. The effect of SES might have confounded the impact of pLI in influencing children’s mental health. Although SES (as indexed by the IMD) and inheritance status were statistically controlled in our analyses, future research incorporating a broader range of SES-related risk and protective factors – particularly those operating at the family level, such as parents’ mental health, parenting style, and family relationships – may help to clarify these associations.

Our analyses further revealed that the inheritance type of CNVs (de novo vs. inherited or unknown) plays a crucial role in predicting the severity of neurodevelopmental and psychiatric outcomes, as well as moderating the effect of pLI. Children with de novo CNVs exhibited lower DQ and higher rates of physical disabilities. This result corroborates and extends the findings by Huguet et al^21^ and Pizzo et al ^34^ on the relationship between CNV inheritance and phenotypes, by showing a direct correlation between the inheritance of a CNV and its effect on severity of developmental delay in children with ID. We also found that children with de novo (versus familial) CNVs had a lower likelihood of psychiatric conditions, which echoes a previous investigation within the IMAGINE-ID that found higher rates of emotional, behavioural, hyperactivity, and autistic diagnoses within the familial group.^14^

Additionally, we find CNV inheritance to moderate the effect of pLI, with a significant prediction of pLI on psychiatric, ADHD, and ASD outcomes in the inherited/unknown groups (only) for deletion CNVs. One possible explanation is that the effect of inherited CNVs may be exerted through indirect genetic pathways, whereby parental carriers of the CNV influence children’s outcomes through shared environment and parent-child interactions. Parents who carry the CNV may themselves experience cognitive or mental health difficulties,^35,36^ which could shape the family environment and indirectly affect children’s MH and ND risk. In contrast, for de novo CNVs, the absence of parental carriage of the CNV may therefore reduce environmentally mediated effects and corresponding mental health risks, potentially explaining why pLI associations with MH outcomes were not observed in the de novo group. An alternative possibility is that, in the de novo group, the effects of CNVs may be more strongly reflected in cognitive abilities not captured by DQ, which could overshadow psychiatric symptoms within this cohort. Although these mechanisms cannot be directly tested in the present data, they provide plausible explanations for the observed moderation by CNV inheritance.

The unexpected directions of effects within our main results led us to consider whether effects could be non-linear, resulting in a “threshold effect” of summed pLI in predicting neurodevelopmental and mental health outcomes. We found preliminary evidence that this is the case: among children with single deletions, the direction of association between pLI and outcomes reversed depending on the pLI range. Within the lower-range pLI group, increasing pLI was associated with heightened neurodevelopmental and mental health difficulties, whereas in the higher-range pLI group, higher pLI was linked to reduced mental health risk.

A similar threshold-like pattern emerged in relation to ID/DD severity within duplications: pLI was positively associated with higher DQ (less severe delay) in the lower-pLI sub-group, but more severe physical disabilities and lower DQ in the higher-pLI group. These findings again highlight the very complex and context-dependent nature of the relationship between pLI, MH and ND outcomes.

The reasons for such a non-linear effect are unclear, but it may reflect the same underlying processes discussed above, including the severity of global impairment that alters the expression or recognition of co-occurring psychiatric symptoms. Similar threshold-dependent or reversed genetic effects have been observed in other studies. For instance, Schmilovich et al^37^ found that pLI and polygenic risk for intelligence showed opposing associations with ASD risk and cognitive outcomes, and that effect of polygenic risk was attenuated in the presence of CNVs with high risks. Similarly, Yeo et al^38^ reported an “antagonistic epistasis” effect in schizophrenia, whereby the negative effect of high CNV deletion burden and SNP-based genetic risk on executive function was attenuated when both risk indices were high. These align with our findings that pLI effects on ND and MH outcomes varied by pLI range, suggesting that accumulated genetic variation exerts non-linear, antagonistic and complex influences on different clinical outcomes.

## Limitations

This study has several limitations. Firstly, the cross-sectional nature of the data limits our ability to infer causal relationships between CNVs and developmental features. Moreover, we used the DAWBA psychiatric disorder likelihood score to measure ND and MH outcomes, which were interpreted as dimensional indicators of psychiatric likelihood. Although it has demonstrated acceptable reliability in neurodevelopmental cohorts, there is limited evidence regarding their sensitivity for capturing specific psychiatric disorders in individuals with severe ID. Another limitation is that our analyses focused primarily on pLI; however, other CNV metrics may capture distinct aspects of genomic risk. . In addition, variation in CNV reporting practices across centres may have influenced the identification of individuals carrying multiple CNVs. Moreover, although SES was included as a covariate, residual confounding from unmeasured family and environmental factors remains possible. Finally, some subgroup analyses were based on relatively small and uneven sample sizes, limiting statistical power.

## Conclusions

Our study’s findings have several implications for both theoretical advances and clinical practice. Exploring the effect of pLI scores and CNV inheritance could help us better understand the complex interplay between genomic and non-genomic factors and their associations with children’s neurodevelopmental and psychiatric outcomes. Practically, it could also improve the precision of diagnosis and targeted support for children with ID, by identifying genetic characteristics that are associated with increased risk of co-occurring neurodevelopmental or mental health problems. Future research could aim to explore the longitudinal pathways linking specific CNV profiles to neurodevelopmental and psychiatric outcomes, to understand how these domains develop over time.

## Acknowledgements

The authors thank the families who participated in the IMAGINE-ID project, and the clinicians and support organisations who facilitated their recruitment. IMAGINE-ID was supported by a grant from the UK Medical Research Council and the UK Medical Research Foundation (MR/L011166/1 and MR/N022572/1). AAB is supported by NIH R01MH133843 and R01MH134896. SASN was supported by the Wellcome Trust (Early Career Award 226392/Z/22/Z). ZC is funded by the Cambridge Trust International CSC scholarship. KB is funded by the UK Medical Research Council (MC_UU_00030/3). All research in the Department of Psychiatry at Cambridge University benefits from the National Institute for Health Research (NIHR) Cambridge Biomedical Research Centre. The views and opinions expressed therein are those of the authors and do not necessarily reflect those of the NIHR, NHS or the Department of Health and Social Care.

## Conflict of Interest / Disclosure

Disclosure: Dr. Alexander-Bloch has worked as a consultant for Octave Bioscience and Centile Bioscience and holds equity in Centile Bioscience. All other authors report no biomedical financial interests or potential conflicts of interest.

## Data Availability Statement

Data from the IMAGINE-ID study are available upon reasonable request and subject to approval by the IMAGINE-ID data access committee and relevant governance procedures.

## Notes

### Author Declarations

The London-Queen Square Research Ethics Committee gave ethical approval for this work.

