## Supplementary materials for "Are CNV Risk Scores Linked to Neurodevelopmental and Mental Health Characteristics Within CNV-Associated Intellectual Disability?"

### Supplementary Methods

#### Supplementary Method 1. CNV risk score calculation

CNVs that were recorded not in the hg19 build (i.e., hg15, hg16, hg17, hg18, hg38) were lifted over into hg19 space, using liftover chains obtained from the UCSC genome browser, to allow for consistent gene mappings. The gene-level risk score data for pLI, LOEUF, pHaplo and pTriplo were downloaded from previous papers (Collins et al., 2022; Karczewski et al., 2020). The CNV data in hg19 were then converted to GRanges objects, to identify overlaps with the risk score data. For a CNV, the sum pLI score was calculated across the pLI of the genes encompassed. Similarly, a cumulative inverse LOEUF score was calculated as the sum of the inverse LOEUF of all involved genes. The pHaplo and pTriplo scores were calculated separately for deletions and duplications. In the main analysis with single CNV cases, we calculated the risk scores separately for deletions and duplications. In the extended analysis which also contains multiple CNV cases, we calculate the risk scores at participant level as the sum score of all CNVs for each CNV metric.

### Supplementary Figures

#### Figure S1. CNV risk score calculation process

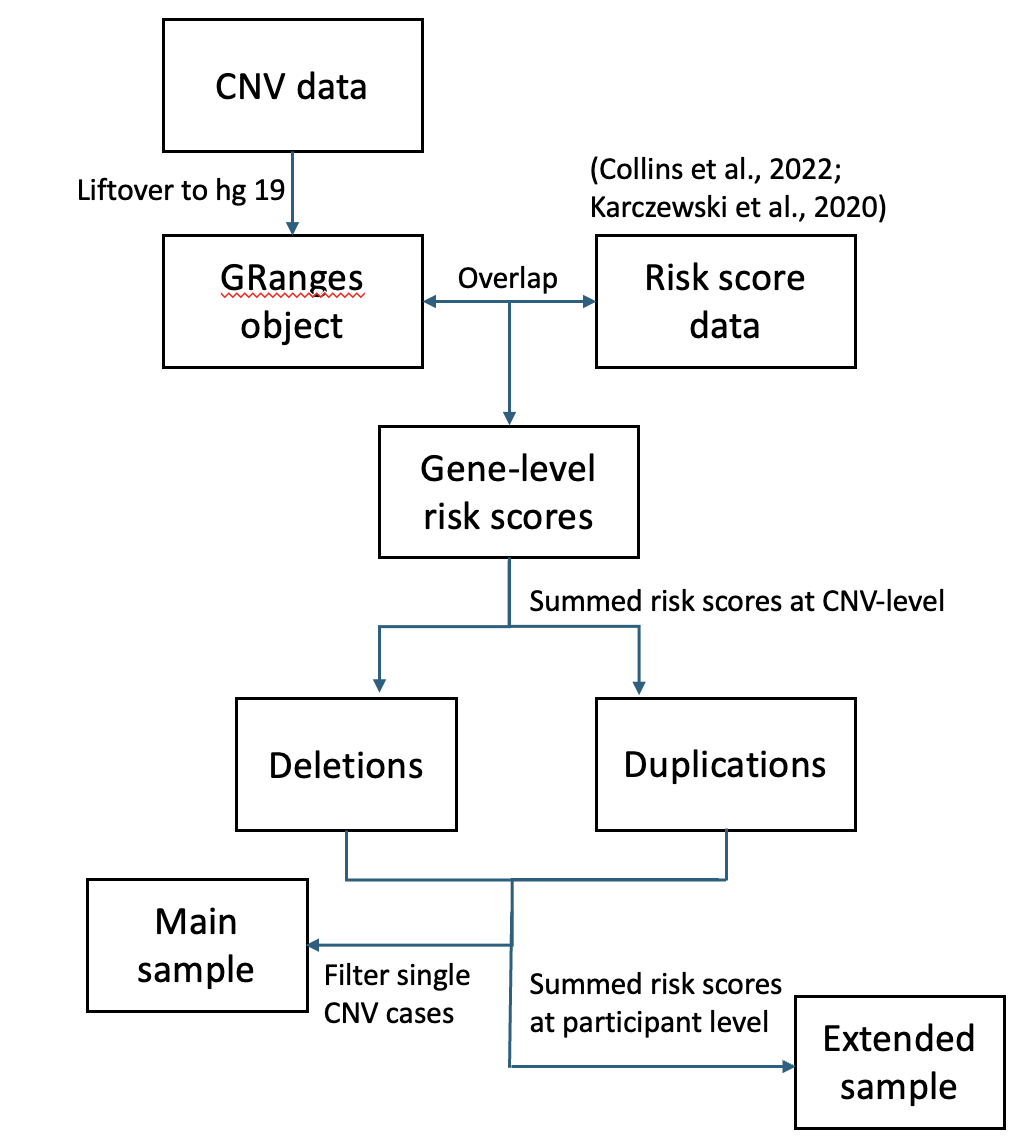

Figure S1. CNV risk score calculation process for the main analysis and extended analysis

#### Figure S2. Distribution of pLI

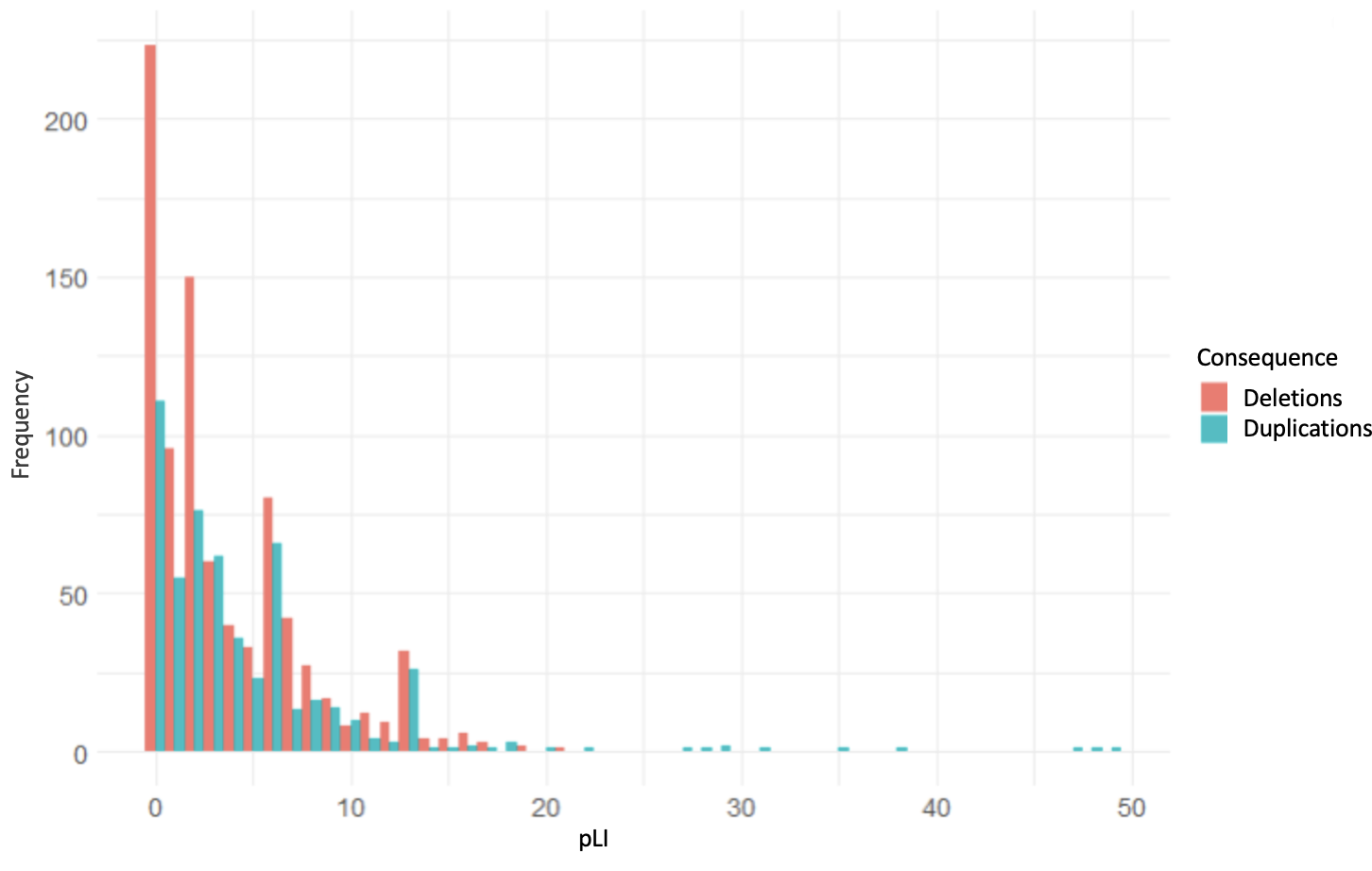

Figure S2. Distributions of (single CNV) pLI scores within Sample 1.

#### Figure S3. Correlation Matrix

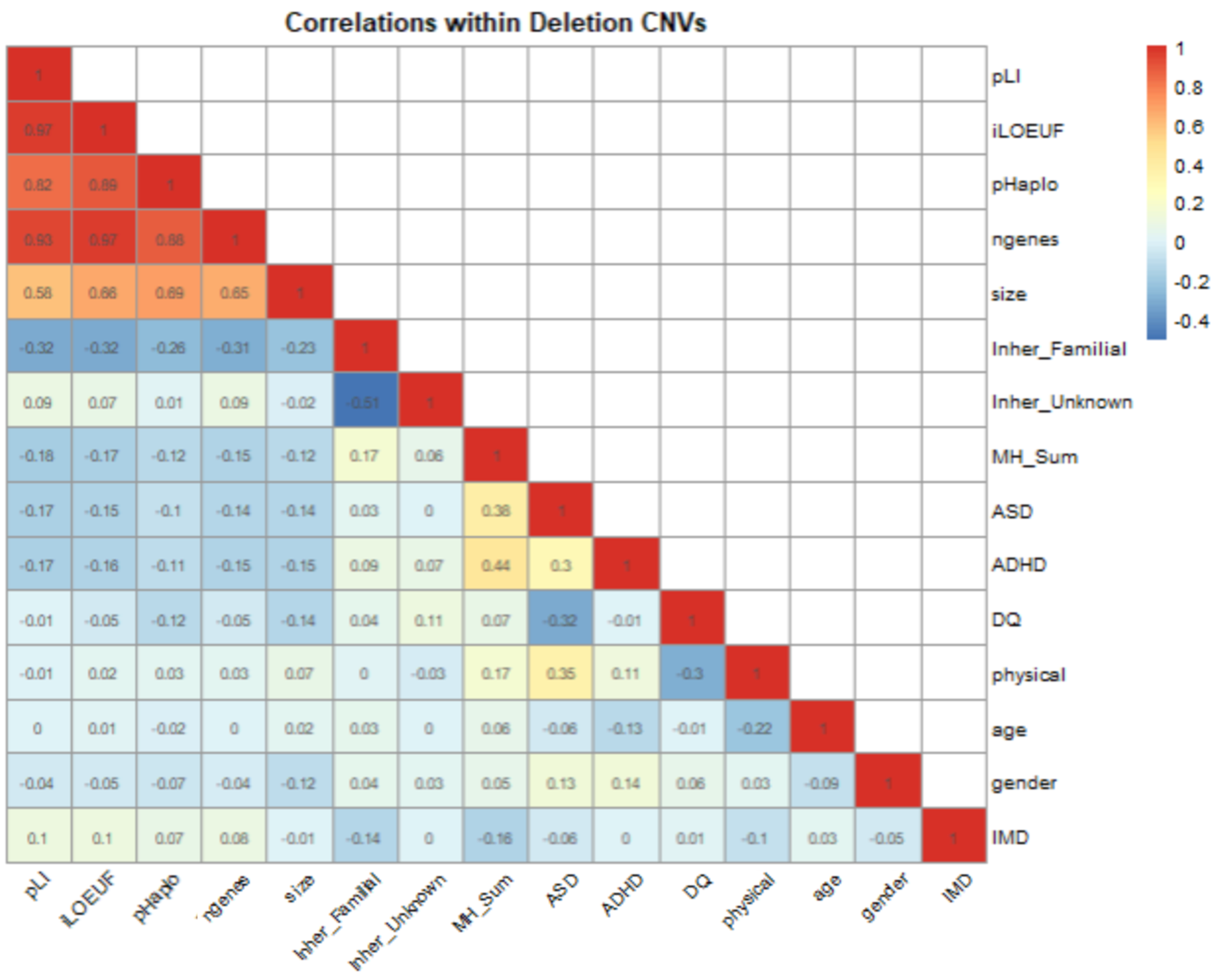

Figure S3a. Correlation matrix in deletions

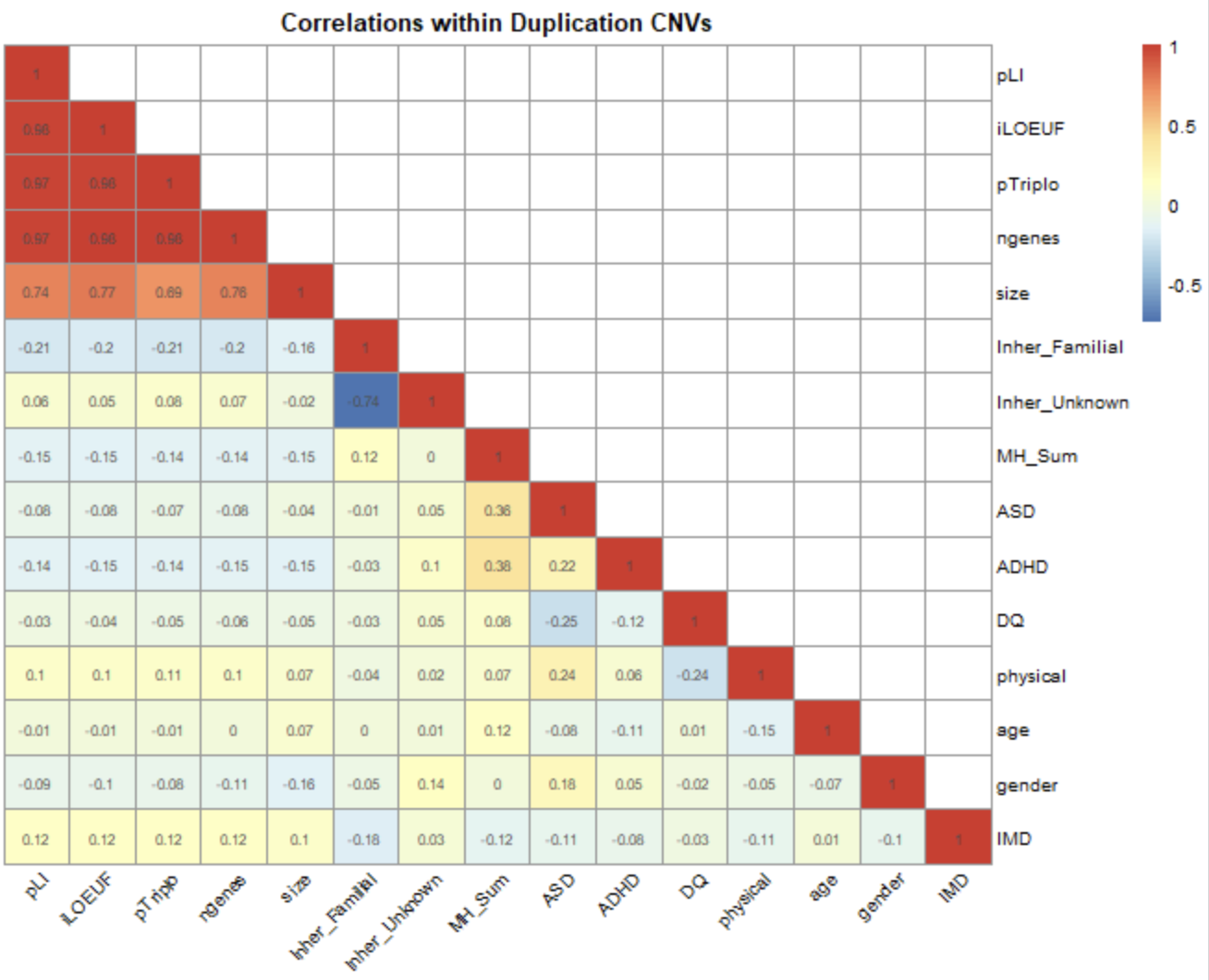

Figure S3b. Correlation matrix in duplications

### Supplementary Tables

| Table S1. Number of participants with different deletion and duplication combinations | | | | | | |
| --- | --- | --- | --- | --- | --- | --- |
| N Deletions\  N Duplications | **0** | **1** | **2** | **3** | **4** | **5** |
| 0 | 0 | 535 | 46 | 4 | 1 | 1 |
| 1 | 849 | 135 | 16 | 1 | 0 | 0 |
| 2 | 38 | 10 | 1 | 0 | 0 | 0 |
| 3 | 2 | 0 | 0 | 0 | 0 | 0 |
| 6 | 0 | 1 | 0 | 0 | 0 | 0 |

| Table S2. DAWBA psychiatric conditions | | |
| --- | --- | --- |
|  | **Item** | **Multi-informant ratings** |
| PsyLi_sum  (Mental Health  Sum Score) | Separation anxiety | 0-5 |
|  | Specific phobia | 0-4 |
|  | Social phobia | 0-4 |
|  | OCD | 0-4 |
|  | General anxiety | 0-4 |
|  | Depression | 0-5 |
|  | ODD | 0-5 |
|  | Conduct disorder | 0-5 |
|  | Tic | 0-5 |
| Neurodevelopmental traits | ASD | 0-5 |
|  | ADHD | 0-5 |

| Table S3. DAWBA Physical Disability Items | |
| --- | --- |
| Physical Condition | Response Type |
| Bed wetting | Binary (0 = No, 1 = Yes) |
| Soiling pants | Binary (0 = No, 1 = Yes) |
| Convulsion/seizures/epilepsy | Binary (0 = No, 1 = Yes) |
| Severe speech or language problems | Binary (0 = No, 1 = Yes) |
| Severe visual impairment or blindness | Binary (0 = No, 1 = Yes) |
| Severe hearing impairment or deafness | Binary (0 = No, 1 = Yes) |
| Severe problems with movement or co-ordination (e.g., cerebral palsy/dyspraxia) | Binary (0 = No, 1 = Yes) |
| Other severe illness or disability | Binary (0 = No, 1 = Yes) |

| Table S4. Recurrent CNVs in individuals with a single CNV | |
| --- | --- |
| CNV | **Frequency** |
| 1q21.1 deletion | 32 |
| 2p16.3 deletion | 44 |
| 3q29 deletion | 7 |
| 15q11.2 deletion | 80 |
| 15q13.3 deletion | 5 |
| 16p11.2 deletion | 98 |
| 16p11.2 duplication | 58 |
| 22q11.2 deletion | 48 |
| 22q11.2 duplication | 43 |

| Table S5. Univariate models of different CNV risk score metrics predicting MH and ND outcomes | | | | | | | | |
| --- | --- | --- | --- | --- | --- | --- | --- | --- |
| Predictors | **Outcomes** | **Conseq** | **std β** | **95%CI lower** | **upper** | **p value** | **adjusted r2** | **AIC** |
| pLI | MH_Sum | deletions | -0.19 | -0.25 | -0.09 | <. 001 | 0.05 | 3632.46 |
|  |  | duplications | -0.16 | -0.23 | -0.03 | 0.009 | 0.04 | 2527.71 |
|  | ADHD | deletions | -0.05 | -0.21 | -0.06 | < .001 | 0.03 | 2506.38 |
|  |  | duplications | -0.02 | -0.17 | 0.01 | 0.09 | 0.02 | 1592.3 |
|  | ASD | deletions | -0.05 | -0.18 | -0.04 | 0.002 | 0.04 | 2754.68 |
|  |  | duplications | -0.004 | -0.11 | 0.07 | 0.72 | 0.04 | 1776.02 |
|  | DQ | deletions | -.0002 | -0.08 | 0.07 | 0.93 | <.001 | -156.71 |
|  |  | duplications | -.002 | -0.18 | 0.03 | 0.15 | 0.001 | -118.61 |
|  | Physical disabilities | deletions | 0.03 | -0.02 | 0.13 | 0.16 | 0.04 | 2731.55 |
|  |  | duplications | 0.02 | -0.02 | 0.16 | 0.14 | 0.02 | 1768.16 |
| pHaplo  pTriplo | MH_Sum | deletions | -0.12 | -0.20 | -0.04 | 0.01 | 0.03 | 3641.72 |
|  |  | duplications | -0.08 | -0.22 | -0.02 | 0.02 | 0.04 | 2528.47 |
|  | ADHD | deletions | -0.01 | -0.14 | 0.01 | 0.1 | 0.02 | 2516.14 |
|  |  | duplications | -0.01 | -0.18 | 0.01 | 0.07 | 0.02 | 1591.79 |
|  | ASD | deletions | -0.01 | -0.11 | 0.03 | 0.28 | 0.03 | 2872.24 |
|  |  | duplications | -0.01 | -0.1 | 0.08 | 0.79 | 0.04 | 1805.62 |
|  | DQ | deletions | -0.07 | -0.14 | 0.004 | 0.066 | 0.027 | 18.7 |
|  |  | duplications | -.003 | -0.10 | 0.10 | 0.95 | <.001 | 78.33 |
|  | Physical disabilities | deletions | 0.08 | 0.001 | 0.15 | 0.05 | 0.03 | 2572.23 |
|  |  | duplications | 0.06 | -0.03 | 0.15 | 0.22 | 0.03 | 1645.59 |
| CNV size | MH_Sum | deletions | -0.14 | -0.22 | -0.05 | < .001 | 0.04 | 3888.25 |
|  |  | duplications | 0.05 | -0.05 | 0.15 | 0.328 | 0.02 | 2680.36 |
|  | ADHD | deletions | -.052 | -0.12 | 0.02 | 0.16 | 0.04 | 2605.95 |
|  |  | duplications | 0.03 | -0.06 | 0.19 | 0.56 | 0.02 | 1630.55 |
|  | ASD | deletions | -0.06 | -0.13 | 0.01 | 0.12 | 0.03 | 2870.98 |
|  |  | duplications | 0.04 | -0.04 | 0.13 | 0.33 | 0.04 | 1804.74 |
|  | DQ | deletions | -0.12 | -0.19 | -0.05 | 0.002 | 0.04 | 12.3 |
|  |  | duplications | -.15 | -0.25 | -0.05 | 0.003 | 0.01 | 69.55 |
|  | Physical disabilities | deletions | 0.12 | 0.04 | 0.19 | 0.002 | 0.04 | 2566.75 |
|  |  | duplications | 0.06 | -0.02 | 0.02 | 0.17 | 0.04 | 1645.25 |
| Note: significant effects are shown in bold. Demographic covariates included in all models are age, gender and IMD. | | | | | | | | |

| Table S6 Model fit indices from main analysis | | | | | | | | | |
| --- | --- | --- | --- | --- | --- | --- | --- | --- | --- |
|  |  | Demographic covariate | | + pLI | | + inheritance | | + pLI + inheritance | |
| DV | **Conseq** | **adjusted r2** | **AIC** | **adjusted r2** | **AIC** | **adjusted r2** | **AIC** | **adjusted r2** | **AIC** |
| MH_Sum | deletions | 0.019 | 3897.55 | 0.054 | 3878.46 | 0.059 | 3876.18 | **0.078** | **3865.90** |
|  | duplications | 0.022 | 2679.33 | 0.040 | 2674.12 | 0.029 | 2679.52 | **0.040** | **2675.92** |
| ADHD | deletions | 0.025 | 2605.92 | 0.040 | 2595.82 | 0.039 | 2597.57 | **0.048** | **2591.28** |
|  | duplications | 0.022 | 1628.89 | 0.025 | 1628.51 | 0.037 | 1623.68 | **0.041** | **1622.46** |
| ASD | deletions | 0.028 | 2871.43 | 0.039 | 2863.67 | 0.026 | 2875.01 | **0.040** | **2864.73** |
|  | duplications | **0.040** | **1803.69** | 0.04 | 1805.61 | 0.040 | 1805.72 | 0.038 | 1807.58 |
| DQ | deletions | 0.023 | 20.10 | 0.022 | 2575.74 | 0.040 | 10.39 | **0.043** | **9.44** |
|  | duplications | **<.001** | **76.34** | <.001 | 78.3 | <.001 | 79.23 | <.001 | 81.21 |
| Physical disabilities | deletions | 0.023 | 2574.15 | 0.022 | 2575.74 | 0.030 | 2571.31 | **0.029** | **2573.29** |
|  | duplications | **0.033** | **1645.12** | 0.034 | 1645.47 | 0.033 | 1646.66 | 0.033 | 1647.74 |
| Note: model comparisons are conducted based on the adjusted r^2^ and the AIC scores, for the different dependent variables (DV) and combining different predictors. Model with the best fit for each DV is shown in bold. | | | | | | | | | |

| Table S7. Model estimates of regressions by inheritance group | | | | | |
| --- | --- | --- | --- | --- | --- |
| DV | Conseq |  | IV | std β | p value |
| MH_Sum | Deletion | De Novo | pLI | 0.06 | 0.46 |
|  |  | Inherited | **pLI** | **-0.20** | **0.01** |
|  |  | Unknown | **pLI** | **-0.22** | **< .001** |
|  | Duplication | De Novo | pLI | -0.07 | 0.6 |
|  |  | Inherited | pLI | -0.12 | 0.11 |
|  |  | Unknown | pLI | -0.12 | 0.13 |
| ADHD | Deletion | De Novo | pLI | -0.04 | 0.5 |
|  |  | Inherited | **pLI** | **-0.29** | **<< .001** |
|  |  | Unknown | pLI | -0.12 | 0.05 |
|  | Duplication | De Novo | pLI | -0.14 | 0.26 |
|  |  | Inherited | pLI | 0.01 | 0.89 |
|  |  | Unknown | pLI | -0.11 | 0.16 |
| ASD | Deletion | De Novo | pLI | -0.1 | 0.13 |
|  |  | Inherited | **pLI** | **-0.15** | **0.03** |
|  |  | Unknown | **pLI** | **-0.15** | **0.007** |
|  | Duplication | De Novo | pLI | 0.06 | 0.6 |
|  |  | Inherited | pLI | -0.1 | 0.11 |
|  |  | Unknown | pLI | 0.05 | 0.51 |
| Physical Disability | Deletion | De Novo | pLI | 0.08 | 0.27 |
|  |  | Inherited | pLI | 0.04 | 0.59 |
|  |  | Unknown | pLI | -0.01 | 0.92 |
|  | Duplication | De Novo | pLI | -0.06 | 0.63 |
|  |  | Inherited | pLI | 0.06 | 0.35 |
|  |  | Unknown | pLI | 0.14 | 0.06 |
| DQ | Deletion | De Novo | pLI | -0.04 | 0.57 |
|  |  | Inherited | **pLI** | **0.16** | **0.05** |
|  |  | Unknown | pLI | 0.08 | 0.19 |
|  | Duplication | De Novo | pLI | -0.18 | 0.19 |
|  |  | Inherited | pLI | -0.01 | 0.94 |
|  |  | Unknown | pLI | -0.13 | 0.1 |
| Note: significant effects are shown in bold. DV = dependent variable, IV = independent variable. In the deletion group (N = 849), inheritance subgroups comprised 268 de novo, 232 inherited, and 349 unknown cases. In the duplication group (N = 535), inheritance subgroups comprised 80 de novo, 249 inherited, and 206 unknown cases. | | | | | |

| Table S8. Regression models in lower pLI and higher pLI sub-groups | | | | | | | | | | | | |
| --- | --- | --- | --- | --- | --- | --- | --- | --- | --- | --- | --- | --- |
| Conseq | pLI range | IV | PsyLi_sum | p | ASD | p | ADHD | p | DQ | p | Physical | p |
| Deletions | pLI <= 1.18  (n=261) | pLI | **0.18 (0.04, 0.31)** | **0.01** | 0.02 (-0.11, 0.15) | 0.77 | 0.06 (-0.07, 0.20) | 0.34 | 0.12 (-0.02, 0.27) | 0.08 | 0.08 (-0.06, 0.22) | 0.23 |
|  |  | inher_fa | **0.43 (0.53, 1.21)** | **<.001** | -0.09 (-0.52, 0.14) | 0.25 | **0.25 (0.17, 0.84)** | **0.002** | **0.21 (0.06, 0.79)** | **0.02** | -0.14 (-0.64, 0.06) | 0.1 |
|  |  | inher_un | **0.44 (0.57, 1.28)** | **<.001** | -0.01 (-0.36, 0.32) | 0.91 | **0.18 (0.04, 0.73)** | **0.02** | 0.17 (-0.02, 0.72) | 0.06 | -0.15 (-0.69, 0.05) | 0.08 |
|  | pLI > 1.18 (n=588) | pLI | **-0.13 (-0.24, -0.03)** | **0.01** | -0.07 (-0.17, 0.02) | 0.12 | **-0.13 (-0.22, -0.03)** | **0.008** | -0.09 (-0.19, 0.01) | 0.07 | 0.05 (-0.05, 0.15) | 0.36 |
|  |  | inher_fa | 0.11 (-0.03, 0.53) | 0.08 | -0.04 (-0.34, 0.17) | 0.51 | 0.02 (-0.22, 0.29) | 0.77 | 0.10 (-0.02, 0.53) | 0.06 | -0.06 (-0.38, 0.15) | 0.39 |
|  |  | inher_un | 0.08 (-0.08, 0.38) | 0.18 | 0.01 (-0.19, 0.21) | 0.91 | **0.11 (0.01, 0.42)** | **0.04** | **0.13 (0.05, 0.47)** | **0.02** | -0.08 (-0.37, 0.06) | 0.14 |
| Duplications | pLI <= 1.18 (n=126) | pLI | -0.04 (-0.25, 0.17) | 0.68 | -0.02 (-0.20, 0.16) | 0.84 | 0.01 (-0.19, 0.21) | 0.94 | **0.24 (0.04, 0.44)** | **0.01** | 0.07 (-0.13, 0.27) | 0.47 |
|  |  | inher_fa | 0.14 (-0.43, 1.00) | 0.41 | **-0.32 (-1.29, -0.05)** | **0.03** | -0.08 (-0.82, 0.50) | 0.62 | **0.57 (0.50, 1.83)** | **<.001** | -0.02 (-0.69, 0.60) | 0.89 |
|  |  | inher_un | 0.04 (-0.86, 0.67) | 0.8 | **-0.33 (-1.45, -0.10)** | **0.02** | 0.05 (-0.60, 0.84) | 0.74 | **0.54 (0.51, 1.99)** | **0.001** | -0.07 (-0.86, 0.55) | 0.66 |
|  | pLI > 1.18 (n=409) | pLI | **-0.14 (-0.25, -0.02)** | **0.02** | 0.01 (-0.10, 0.11) | 0.88 | **-0.11 (-0.22, -0.00)** | **0.04** | **-0.18 (-0.30, -0.06)** | **0.003** | **0.12 (0.01, 0.23)** | **0.03** |
|  |  | inher_fa | 0.13 (-0.08, 0.64) | 0.12 | 0.10 (-0.11, 0.53) | 0.2 | 0.08 (-0.17, 0.49) | 0.33 | -0.16 (-0.68, 0.04) | 0.09 | -0.00 (-0.34, 0.34) | 0.98 |
|  |  | inher_un | 0.13 (-0.08, 0.63) | 0.12 | **0.20 (0.09, 0.71)** | **0.01** | **0.19 (0.07, 0.72)** | **0.02** | -0.09 (-0.53, 0.18) | 0.32 | 0.02 (-0.29, 0.38) | 0.79 |
| Note. Values are standardised regression coefficients (β) with 95% confidence intervals in parentheses. De novo inheritance is the reference group (inher_fa = familial vs. de novo; inher_un = unknown vs. de novo). All models were adjusted for age, sex, and socioeconomic status (IMD decile). pLI sub-groups were defined using a cut-off of 1.18. Higher DQ indicates more age-appropriate functioning; higher Physical Disability scores indicate greater impairment. | | | | | | | | | | | | |

| Table S9. Comparison Between Higher Range and Lower Range pLI groups | | | | |
| --- | --- | --- | --- | --- |
| Variable/Group | **Deletion low** **pLI** | **Deletion high pLI** | **Duplication low pLI** | **Duplication high pLI** |
| N | 261 | 588 | 126 | 409 |
| age (mean, SD) | 8.61 (3.64) | 8.35 (3.71) | 8.97 (3.92) | 8.55 (3.52) |
| PsyLi_adhd (mean, SD) | 3.23 (1.49) | 3.01 (1.49) | 3.21 (1.41) | 3.31 (1.40) |
| PsyLi_sa (mean, SD) | 14.34 (6.77) | 12.75 (6.10) | 15.88 (8.03) | 14.95 (6.46) |
| PsyLi_asd (mean, SD) | 3.14 (1.55) | 2.74 (1.65) | 3.10 (1.60) | 2.91 (1.60) |
| DQ (mean, SD) | 0.55 (0.22) | 0.60 (0.21) | 0.58 (0.21) | 0.63 (0.20) |
| Physical (mean, SD) | 2.88 (2.00) | 3.07 (2.06) | 2.56 (1.77) | 2.42 (1.73) |
| IMD decile (top level, %) | 5.41 (2.9) | 5.33 (2.88) | 4.98 (2.94) | 4.74 (2.78) |

| Table S10. Model fit indices from extended analyses | | | | | | | | | | | | | | | | |
| --- | --- | --- | --- | --- | --- | --- | --- | --- | --- | --- | --- | --- | --- | --- | --- | --- |
|  | **Demographic covariate** | | **+ pLI** | | **+ inheritance** | | **+ n Dels + nDups** | | **+ pLI + inheritance** | | **+pLI**  **+ nDels + nDups** | | **+ inheritance**  **+ nDels + nDups** | | **+ pLI + inher**  **+ nDels + nDups** | |
|  | **adjusted r2** | **AIC** | **adjusted r2** | **AIC** | **adjusted r2** | **AIC** | **adjusted r2** | **AIC** | **adjusted r2** | **AIC** | **adjusted r2** | **AIC** | **adjusted r2** | **AIC** | **adjusted r2** | **AIC** |
| MH_Sum | 0.036 | 7392.68 | 0.053 | 7374.67 | 0.073 | 7349.9 | 0.046 | 7382.82 | 0.082 | 7339.99 | 0.066 | 7360.59 | 0.078 | 7346.56 | **0.09** | **7333.12** |
| ADHD | 0.016 | 4912.73 | 0.024 | 4902.14 | 0.030 | 4894.42 | 0.017 | 4913.24 | 0.034 | 4888.53 | 0.027 | 4900.44 | 0.029 | 4897.08 | **0.035** | **4889.46** |
| ASD | **0.031** | **5364.69** | 0.032 | 5364.77 | 0.031 | 5366.18 | 0.030 | 5368.41 | 0.032 | 5366.56 | 0.031 | 5368.23 | 0.030 | 5370.01 | 0.030 | 5370.11 |
| DQ | <.001 | -349.68 | 0.003 | -353.15 | 0.008 | -359.41 | <.001 | -347.87 | **0.009** | **-360.45** | 0.003 | -351.89 | 0.007 | -356.28 | 0.009 | -357.70 |
| Physical  disabilities | 0.022 | 5048.37 | 0.028 | 5041.62 | 0.033 | 5034.78 | 0.046 | 5016.76 | 0.036 | 5031.73 | 0.053 | 5009.57 | 0.053 | 5009.77 | **0.057** | **5005.64** |
| Note: models were compared based on the adjusted r^2^ and AIC indices. The model with the best fit for each outcome was shown in bold. Inher = Inheritance type, nDels/nDups = Total number of deletion and duplication CNVs. | | | | | | | | | | | | | | | | |

| Table S11. Model estimates from extended analyses | | | | | |
| --- | --- | --- | --- | --- | --- |
| DV | **IV** | **std β** | **95%CI lower** | **upper** | **p value** |
| MH_Sum | pLI | **-0.12** | **-0.18** | **-0.06** | **< .001** |
|  | Any_DN | **-0.17** | **-0.22** | **-0.11** | **< .001** |
|  | nDels | -0.03 | -0.11 | 0.04 | 0.42 |
|  | nDups | 0.07 | -0.001 | 0.15 | 0.056 |
| ADHD | pLI | **-0.08** | **-0.14** | **-0.03** | **0.002** |
|  | Any_DN | **-0.1** | **-0.16** | **-0.05** | **< .001** |
|  | nDels | -0.001 | -0.07 | 0.07 | 0.98 |
|  | nDups | 0.05 | -0.02 | 0.12 | 0.19 |
| ASD | n/a |  |  |  |  |
| DQ | pLI | -0.05 | -0.11 | 0.01 | 0.08 |
|  | Any_DN | **-0.09** | **-0.15** | **-0.03** | **0.002** |
| Physical disabilities | pLI | **0.07** | **0.01** | **0.13** | **0.014** |
|  | Any_DN | **0.07** | **0.01** | **0.13** | **0.015** |
|  | nDels | **0.16** | **0.08** | **0.23** | **< .001** |
|  | nDups | 0.01 | -0.07 | 0.08 | 0.88 |
| Note: significant beta coefficients are shown in bold. DV = dependent variable, IV = independent variable. Any_DN: 1 = if any of the CNV is de novo. nDels/nDups = Total number of deletion and duplication CNVs. ASD models are not presented as inclusion of CNV characteristics did not improve model fit. | | | | | |

| Table S12. Model estimates of regressions by inheritance group in extended analyses | | | | | | | | | |
| --- | --- | --- | --- | --- | --- | --- | --- | --- | --- |
| DV | **Group** | **IV** | **std β** | **p value** | **DV** | **Group** | **IV** | **std β** | **p value** |
| MH_Sum | De Novo | pLI | -0.003 | 0.96 | **ADHD** | De Novo | pLI | -0.04 | 0.45 |
|  |  | nDels | -0.08 | 0.29 |  |  | nDels | 0.001 | 0.99 |
|  |  | nDups | 0.02 | 0.78 |  |  | nDups | 0.07 | 0.32 |
|  | No DN | **pLI** | **-0.16** | **<.001** |  | No DN | **pLI** | **-0.11** | **0.001** |
|  |  | nDels | -0.01 | 0.75 |  |  | nDels | 0.0001 | 0.998 |
|  |  | nDups | 0.09 | 0.05 |  |  | nDups | 0.04 | 0.36 |
| ASD | De Novo | pLI | 0.01 | 0.88 | **DQ** | De Novo | **pLI** | **-0.13** | **0.03** |
|  |  | nDels | -0.03 | 0.70 |  |  | nDels | 0.09 | 0.17 |
|  |  | nDups | -0.04 | 0.55 |  |  | **nDups** | **0.18** | **0.01** |
|  | No DN | pLI | -0.06 | 0.07 |  | No DN | pLI | -0.03 | 0.42 |
|  |  | nDels | 0.01 | 0.81 |  |  | nDels | -0.07 | 0.1 |
|  |  | nDups | 0.03 | 0.44 |  |  | nDups | -0.05 | 0.27 |
| Physical | De Novo | pLI | 0.09 | 0.11 |  |  |  |  |  |
|  |  | nDels | 0.07 | 0.35 |  |  |  |  |  |
|  |  | nDups | -0.09 | 0.21 |  |  |  |  |  |
|  | No DN | **pLI** | **0.06** | **0.05** |  |  |  |  |  |
|  |  | **nDels** | **0.19** | **< .001** |  |  |  |  |  |
|  |  | nDups | 0.04 | 0.39 |  |  |  |  |  |
| Note: significant beta coefficients are shown in bold. DV = dependent variable, IV = independent variable. Any_DN: 1 = if any of the CNV is de novo. nDels/nDups = Total number of deletion and duplication CNVs. | | | | | | | | | |

| Table S13. Regression models in lower range and higher range pLI groups in extended analyses | | | | | | | | | | | |
| --- | --- | --- | --- | --- | --- | --- | --- | --- | --- | --- | --- |
|  |  | **MH_Sum** | | **ASD** | | **ADHD** | | **DQ** | | **Physical** | |
|  | **IV** | **std β** | **p value** | **std β** | **p value** | **std β** | **p value** | **std β** | **p value** | **std β** | **p value** |
| **pLI <= 1.18**  **(n=436)** | pLI | 0.07 | 0.19 | 0.01 | 0.9 | 0.06 | 0.26 | **0.14** | **0.01** | 0.07 | 0.21 |
|  | nDups | 0.06 | 0.49 | -0.01 | 0.88 | 0.07 | 0.38 | 0.07 | 0.39 | -0.02 | 0.78 |
|  | nDels | 0.02 | 0.49 | 0.01 | 0.88 | 0.12 | 0.12 | 0.03 | 0.71 | 0.06 | 0.46 |
|  | Any_DN | **-0.27** | **< .001** | 0.06 | 0.24 | **-0.13** | **0.01** | **-0.18** | **0.001** | 0.1 | 0.07 |
| **pLI > 1.18**  **(n=1210)** | pLI | **-0.12** | **0.007** | 0.005 | 0.89 | **-0.09** | **0.008** | **-0.15** | **< .001** | **0.11** | **0.001** |
|  | nDups | 0.08 | 0.06 | 0.03 | 0.52 | 0.05 | 0.22 | 0.002 | 0.97 | -0.004 | 0.92 |
|  | nDels | -0.06 | 0.20 | -0.01 | 0.89 | -0.03 | 0.42 | -0.04 | 0.37 | **0.17** | **< .001** |
|  | Any_DN | **-0.12** | **< .001** | -0.04 | 0.27 | **-0.09** | **0.006** | -0.04 | 0.24 | 0.04 | 0.28 |
| Note: significant effects are shown in bold. DV = dependent variable, IV = independent variable. Any_DN: 1 = if any of the CNV is de novo. nDels/nDups = Total number of deletion and duplication CNVs**.** | | | | | | | | | | | |
